# Syphilis clustering among young pregnant women in Kampala, Uganda

**DOI:** 10.1101/2025.08.21.25334202

**Authors:** Rogers Nsubuga, Timothy R Muwonge, Andrew Mujugira, Barbara Castelnuvo, Edith Nakku-Joloba, Rosalind Parkes-Ratanshi, Yukari C. Manabe, Agnes Kiragga

## Abstract

**Introduction:** In Uganda, the spatial distribution of syphilis varies by age, gender, and region. Identifying clusters (subsets of administrative subdivisions) with high syphilis prevalence could boost efforts to eliminate mother-to-child transmission of syphilis. We examined spatial variations and clustering of syphilis prevalence among pregnant young women in Central Uganda.

**Methods:** We analysed secondary data from a randomised trial that evaluated the effectiveness of three antenatal syphilis partner notification approaches (NCT02262390). This study analysed clustering of syphilis prevalence by administrative division in Kampala and Wakiso districts, using Moran’s I tests and Local Indicator of Spatial Association (LISA). We used the Kulldorff Spatial-Scan Poisson model to classify divisions with high or low syphilis prevalence (HP/LP) based on 95% statistical significance. We estimated prevalence ratios for sociodemographic and bio-behavioural HIV risk factors associated with clustering, stratified by HIV status, using modified Poisson regression.

**Results:** Of 422 young women diagnosed with syphilis, 26 (6%) had HIV and syphilis. The median age was 26 years (IQR 24-29). Most (314, 74%) were in monogamous marriages, and half (50%) had ≤13 years of schooling. Syphilis prevalence clustering was negatively correlated with being in a polygamous marriage (adjusted prevalence ratio [APR]=0.64; 95%: 0.47-0.88), having an unplanned pregnancy (APR=0.78; 95% CI: 0.64-0.93) and HIV testing >3 months prior (APR=0.83, 95% CI: 0.72-0.95). Syphilis prevalence was significantly higher in 3 of 12 clusters–Kasangati Town Council (Relative Risk [RR]=2.79, p<0.0001), Kawempe (RR=2.52, p<0.0001), and Nabweru (RR=1.95, p=0.0002), and lower in one cluster–Kyengera Town Council (RR=0.12, p<0.0001). Notably, no significant clustering was detected among women with HIV (p>0.05). Random patterns of syphilis prevalence were detected across all divisions (Moran’s I=0.08, p=0.19). However, some neighbouring divisions had similar prevalence: Kawempe (1.06, p=0.02) and Nabweru (0.54, p=0.045). LISA analysis confirmed high syphilis prevalence in northern divisions (Kawempe and Nabweru; p=0.01). By contrast, Central Region had neighbouring low and high prevalence divisions (Kawempe and Central; p=0.001).

**Conclusion:** Syphilis prevalence was similar within neighbouring divisions, but highest in Kasangati Town Council and Kawempe. Scaling up spatial analysis application tools enables the detection of clusters where interventions can be targeted to eliminate congenital syphilis.

## Introduction

In sub-Saharan Africa (SSA), adolescent girls and young women (AGYW) aged 15 to 24 years are at high risk of HIV and other sexually transmitted infections (STIs) [1]. In 2021, AGYW accounted for 27% of all new HIV infections in this region [1]. In SSA, studies indicate higher STI prevalence among younger women (15–24 years) compared with older (25–49 years), and they were three times as likely to acquire HIV compared to their male counterparts [2, 3]. Uganda has the highest HIV prevalence in East Africa; AGYW make up 7.2% of the people living with HIV, with the highest prevalence in the Central Region [4]. Syphilis prevalence was 1.3% among school-going girls [5], 3% among women attending facility antenatal clinics, and was more common among those with HIV (6.2%) than without (1.8%) [6]. Elimination of mother-to-child transmission of syphilis is a global health priority [7]. Thus, identifying high-prevalence clusters in which pregnant women have active syphilis infection is key to preventing adverse birth outcomes [8].

Recent research has increasingly employed spatial analysis to better understand the geographical distribution and clustering of HIV and other STIs, including syphilis. A scoping review demonstrated that mapping these patterns highlighted high-burden areas and helped uncover potential environmental and social determinants influencing disease transmission [9]. Spatial methods have been applied to detect localized HIV clusters in urban settings, revealing overlaps with other STIs and guiding targeted public health interventions [10]. Spatial analysis relies on three general methodological approaches to detect clustering: 1) based on division counts, 2) autocorrelative adjacencies of divisions with high counts, and 3) determining the distance between events [11]. Spatial clustering techniques are essential for statistical consideration and form the initial steps in developing a model predicting disease risk sites [12, 13]. In this study, we examined spatial autocorrelation and clustering of syphilis prevalence among pregnant young women in Central Uganda. We also investigated potential factors associated with syphilis prevalence among AGYW.

## METHODS

### Population and Procedures

The Syphilis Treatment of Partners (STOP) study was a randomised clinical trial that compared the effectiveness of three partner notification strategies after antenatal syphilis screening among ∼17,000 pregnant women in Uganda (www.ClinicalTrials.gov, NCT02262390, 2014-October-08) [14]. This trial took place from 12^th^ February 2015 to 17^th^ February 2016. Women attending antenatal care in Central Uganda were included in the trial if they were aged 18 years or older, had positive pregnancy and treponemal antibody rapid tests, and had a known sexual partner. HIV testing was performed using rapid HIV tests according to national guidelines [15]. Syphilis testing was conducted at enrolment using a point-of-care lateral flow test (SD Bioline, Standard Diagnostics, Inc., Republic of Korea) and confirmed using a lipoidal rapid plasmin reagin (RPR) test (Fortress Diagnostics, United Kingdom). Pregnant women and their male partners received free treatment for syphilis if they tested positive.

### Statistical Analyses

This present analysis utilized STOP trial deidentified data that was accessed on 02^nd^ August 2022. The primary outcome was spatial variation in syphilis prevalence among young pregnant women in Kampala and Wakiso districts. Women from the STOP study were included in the analyses if they were aged 35 years or younger and had a positive syphilis test at baseline. The district administrative division was the unit of analysis. Two spatial autocorrelation methods were used to assess syphilis clustering among AGYW: 1) Global Moran’s I test to measure the overall spatial autocorrelation and detect whether syphilis prevalence rates were spatially clustered, dispersed, or randomly distributed across divisions; 2) Local Indicators of Spatial Association (LISA) to identify and map specific divisions where syphilis prevalence was significantly clustered, highlighting potential high-high (hot spot) or low-low (cold spot) [16]. Neighbouring divisions were determined using the first-order queen polygon continuity method [17].

We used a Kulldorff Spatial-Scan Poisson model to identify divisions with high syphilis prevalence among young women attending antenatal care in Kampala and Wakiso districts (Central Uganda). The number of persons diagnosed with syphilis in a division was assumed to be Poisson-distributed according to an underlying population, young women at risk. The population at risk was determined by extracting from the census report the number of AGYW who were currently or had been in a marital relationship 12 months before enrolment within the respective divisions [18]. The population of AGYW living with HIV was determined by computing the product of the district HIV prevalence [6] and the population of AGYW per division, assuming district HIV prevalence was uniform across different district divisions.

The baseline risk of having multiple sexual partners in the past 12 months was assumed to be 19% for all AGYW and 14% for AGYW living with HIV, based on the Uganda Population-based HIV Impact Assessment [6]. We computed the spatial scan statistic using the Kulldorff Spatial Scan Poisson model. The maximum number of Monte Carlo replications was set to 9,999, and statistical significance was set at p <0.05. Divisions were considered high-prevalence if they had statistically significant spatial scan statistics, i.e., when the log-likelihood ratio (LLR) exceeded the critical value generated from the Monte Carlo simulations. Divisions were classified as either high prevalence or low prevalence (HP/LP) based on the spatial scan statistic set at 5% significance, thus creating a binary outcome variable to indicate syphilis clustering. Using modified Poisson regression, we estimated adjusted prevalence ratios for sociodemographic and bio-behavioural HIV risk factors associated with syphilis clustering, stratified by HIV status based on 5% significance. Data were analysed using SaTScan and STATA 18.

### Ethics Approval

The STOP trial was approved by the Joint Clinical Research Centre Research Ethics Committee (JC1214), the Uganda National Council for Science and Technology (HS1681), and the Johns Hopkins IRB (NA_00012998/CR00015330). All study participants in the STOP trial gave informed written consent for randomization and for temporary specimen storage (blood)[14].

## RESULTS

Of 442 pregnant women with syphilis in the STOP study, 422 were included in the present analysis. The remaining 20 women were excluded because they were either older than 35 years or resided outside the 12 divisions in the study catchment area. Results from show that the analysis included.

The median age was 26 years (interquartile range [IQR]: 24-29). About half (224, 53%) were employed, most (314, 74%) were in monogamous marriages, and half (50%) had completed ≤13 years of schooling (**Table 1**). The majority (378, 90%) reported at least two prior pregnancies, 319 (76%) reported ever experiencing a stillbirth, and 125 (30%) reported having ever had an unintended pregnancy. At enrolment, 252 (60%) women reported HIV testing in the past three months, and 26 (6%) were diagnosed with both HIV and syphilis. Eighty-one male partners (19%) were enrolled in the study; the median age was 30 years (IQR 28.0, 39.0), and almost all 78 (97%) were employed. Of these, 23 (28%) reported having other sexual partners, of whom 78% (18/23) reported more than two partners.

**Table 1:** Participant characteristics across the prevalence clusters.

| Participant characteristics | General<br>(N=422) | Low prevalence<br>(151, 35.8%) | High prevalence<br>(271, 64.2%) |
| --- | --- | --- | --- |
| **Age of participant** |  |  |  |
| Median (Q1, Q3) | 26.0 (24.0, 29.0) | 26.0 (24.0, 29.0) | 26.0 (24.0, 29.0) |
| **Education level** |  |  |  |
| Primary | 178 (42.2%) | 57 (32.0%) | 121 (68.0%) |
| Secondary | 211 (50.0%) | 81 (38.4%) | 130 (61.6%) |
| Tertiary | 33 (7.8%) | 13 (39.4%) | 20 (60.6%) |
| **Marital status<sup>1</sup>** |  |  |  |
| Married monogamous | 314 (74.4%) | 103 (32.8%) | 211 (67.2%) |
| Married polygamous | 90 (21.3%) | 44 (48.9%) | 46 (51.1%) |
| Cohabiting | 18 (4.3%) | 4 (22.2%) | 14 (77.8%) |
| **Undertaken paid work prior 3 months** |  |  |  |
| No | 194 (46.0%) | 74 (38.1%) | 120 (61.9%) |
| Yes | 228 (54.0%) | 77 (33.8%) | 151 (66.2%) |
| **Main employment status** |  |  |  |
| Unemployed/Not working | 198 (46.9%) | 74 (37.4%) | 124 (62.6%) |
| Self-employed/Small business | 192 (45.5%) | 64 (33.3%) | 128 (66.7%) |
| Public/Private employee | 32 (7.6%) | 13 (40.6%) | 19 (59.4%) |
| **Ever had an unintended pregnancy<sup>1</sup>** |  |  |  |
| No | 297 (70.4%) | 91 (30.6%) | 206 (69.4%) |
| Yes | 125 (29.6%) | 60 (48.0%) | 65 (52.0%) |
| **Prior pregnancy outcome<sup>1</sup>** |  |  |  |
| Still births | 319 (75.6%) | 101 (31.7%) | 218 (68.3%) |
| Miscarriage | 60 (14.2%) | 27 (45.0%) | 33 (55.0%) |
| Others | 43 (10.2%) | 23 (53.5%) | 20 (46.5%) |
| **Presence of genital ulcers** | 9 (2.1%) | 4 (44.4%) | 5 (55.6%) |
| **Gravid categories** |  |  |  |
| 1 | 44 (10.4%) | 16 (36.4%) | 28 (63.6%) |
| 2+ | 378 (89.6%) | 135 (35.7%) | 243 (64.3%) |
| **HIV testing history<sup>1</sup>** |  |  |  |
| $\leq 3$ months | 170 (40.3%) | 44 (25.9%) | 126 (74.1%) |
| $> 3$ months | 252 (59.7%) | 107 (42.5%) | 145 (57.5%) |
| **HIV status<sup>2</sup>** |  |  |  |
| Positive | 26 (6.2%) | 14 (53.8%) | 12 (46.2%) |
| Negative | 396 (93.8%) | 137 (34.6%) | 259 (65.4%) |
| **Male Partners** |  |  |  |
| **Age of Partner** |  |  |  |
| Median (Q1, Q3) | 30.0 (28.0, 39.0) | 31.0 (28.0, 37.0) | 30.0 (27.0, 40.0) |
| **Partner employed** | 78 (97.5%) | 30 (38.5%) | 48 (61.5%) |
| **Partner's years of work** |  |  |  |
| Median (Q1, Q3) | 8.0 (4.0, 15.0) | 8.0 (3.0, 10.0) | 9.0 (4.0, 15.0) |
| **Having ≥ 1 sexual partner** | 23 (29.1%) | 7 (30.4%) | 16 (69.6%) |
| **Sexual Partners<sup>3</sup>** |  |  |  |
| 1 partner | 5 (21.7%) | 0 (0.0%) | 5 (100%) |
| ≥2 partners | 18 (78.3%) | 7 (38.9%) | 11 (61.1%) |
| **Partner condom use with** |  |  |  |
| No | 16 (59.3%) | 7 (43.8%) | 9 (56.2%) |
| Yes | 11 (40.7%) | 3 (27.3%) | 8 (72.7%) |
| **Partner syphilis results** |  |  |  |
| Negative | 58 (75.3%) | 22 (37.9%) | 36 (62.1%) |
| Positive | 19 (24.7%) | 8 (42.1%) | 11 (57.9%) |

<sup>1,2,3</sup>Statistically significant at p<0.01, p<0.05 and p<0.2 respectively

Of the 12 divisions where women with syphilis were resident, four (33%) had statistically significant rates of syphilis, classified as High Prevalence (HP) divisions. Among all the women enrolled, 271 (64%) were residents of HP divisions. More women in HP divisions had an HIV test compared to LP divisions (p<0.0001), while more women in HP divisions had negative HIV results (p=0.045).

Multivariable analysis revealed that syphilis prevalence in HP divisions was negatively associated with being in a polygamous relationship, having prior unintended pregnancy, and having done HIV testing beyond three months prior (**Table 2**). Women in polygamous relationships were 36% less likely to be living in HP divisions (adjusted prevalence ratio [APR] = 0.64; 95% confidence interval [CI]: 0.47, 0.88; p=0.005). Similarly, women with a history of an unintended pregnancy were 22% less likely to be residents in HP divisions (APR=0.78, 95% CI: 0.64, 0.93, p=0.007). In comparison, women with long duration (at least three months) since the last HIV test were 17% less likely to live in HP divisions (APR=0.83, 95% CI: 0.72, 0.95, P=0.008). However, women with negative HIV serostatus were 39% as likely to reside in an HP division, but this was not statistically significant (APR=1.39, 95% CI: 0.91, 2.11, p=0.12).

**Table 2:** Factors associated with living in a high prevalence division.

|  | Unadjusted |  | Adjusted |  |
| --- | --- | --- | --- | --- |
|  | Prevalence Ratio (95% CI) | P-value | Prevalence Ratio (95% CI) | P-value |
| **Marital status** |  |  |  |  |
| Cohabiting | Ref | - | Ref | - |
| Married monogamous | 0.86 (0.67, 1.12) | 0.27 | 0.82 (0.64, 1.05) | 0.12 |
| Married polygamous | 0.66 (0.48, 0.90) | 0.01 | 0.64 (0.47, 0.88) | 0.005 |
| **Ever had an unintended pregnancy** |  |  |  |  |
| No | Ref | - | Ref | - |
| Yes | 0.75 (0.62, 0.90) | 0.002 | 0.78 (0.64, 0.93) | 0.007 |
| **HIV testing history** |  |  |  |  |
| ≤3 months | Ref | - | Ref | - |
| >3 months | 0.78 (0.68, 0.89) | <0.001 | 0.83 (0.72, 0.95) | 0.008 |
| **HIV status** |  |  |  |  |
| Positive | Ref | - | Ref | - |
| Negative | 1.42 (0.93, 2.16) | 0.11 | 1.39 (0.91, 2.11) | 0.12 |

Using the first-order queen polygon, each division had neighbouring divisions in the range of 1-5, and the mean number of neighbours was 2.5. Global Moran’s I revealed a weak tendency towards syphilis clustering that was not statistically significant (Moran’s I=0.08, p=0.19), suggesting syphilis was randomly distributed across the divisions of Kampala and Wakiso districts. Local Moran’s matrix found that most neighbouring divisions had similar prevalence, except for Kawempe division (I=1.06, p=0.019), which neighboured Nabweru division in the north (I=0.54, p=0.045) (**Table 3**).

**Table 3:** Local Moran’s I test.

| Division | Moran Statistic | P-value | LISA classes |
| --- | --- | --- | --- |
| Central Division | -0.64 | 0.100 | Low-High |
| Gombe Division | -0.65 | 0.236 | Low-High |
| Kajjansi Town Council | 0.42 | 0.343 | Low-High |
| Kasangati Town Council | 0.51 | 0.193 | High-High |
| Kawempe Division | 1.06 | 0.019 | High-High |
| Kira Division | -0.85 | 0.254 | Low-High |
| Kyengera Town Council | 0.21 | 0.434 | Low-Low |
| Makindye Division | 0.02 | 0.898 | Low-Low |
| Nabweru Division | 0.54 | 0.045 | High-High |
| Ndejje Division | 0.28 | 0.255 | Low-Low |
| Rubaga Division | 0.02 | 0.799 | High-High |
| Wakiso Division | 0.06 | 0.819 | Low-Low |

Syphilis prevalence was highest in Kawempe Division and its neighbouring divisions: Rubaga in the south, and Nabweru and Kasangati Town Council in the north (**Figure 1B)**. LISA analysis revealed that high syphilis prevalence and neighbouring divisions in the north were statistically related (Kawempe and Nabweru, p=0.05). Furthermore, the low prevalence in the Central division and the high prevalence in Kawempe division in the south were statistically associated (Central and Kawempe divisions, p=0.001) (**Figure 1C**).

**Figure 1.**
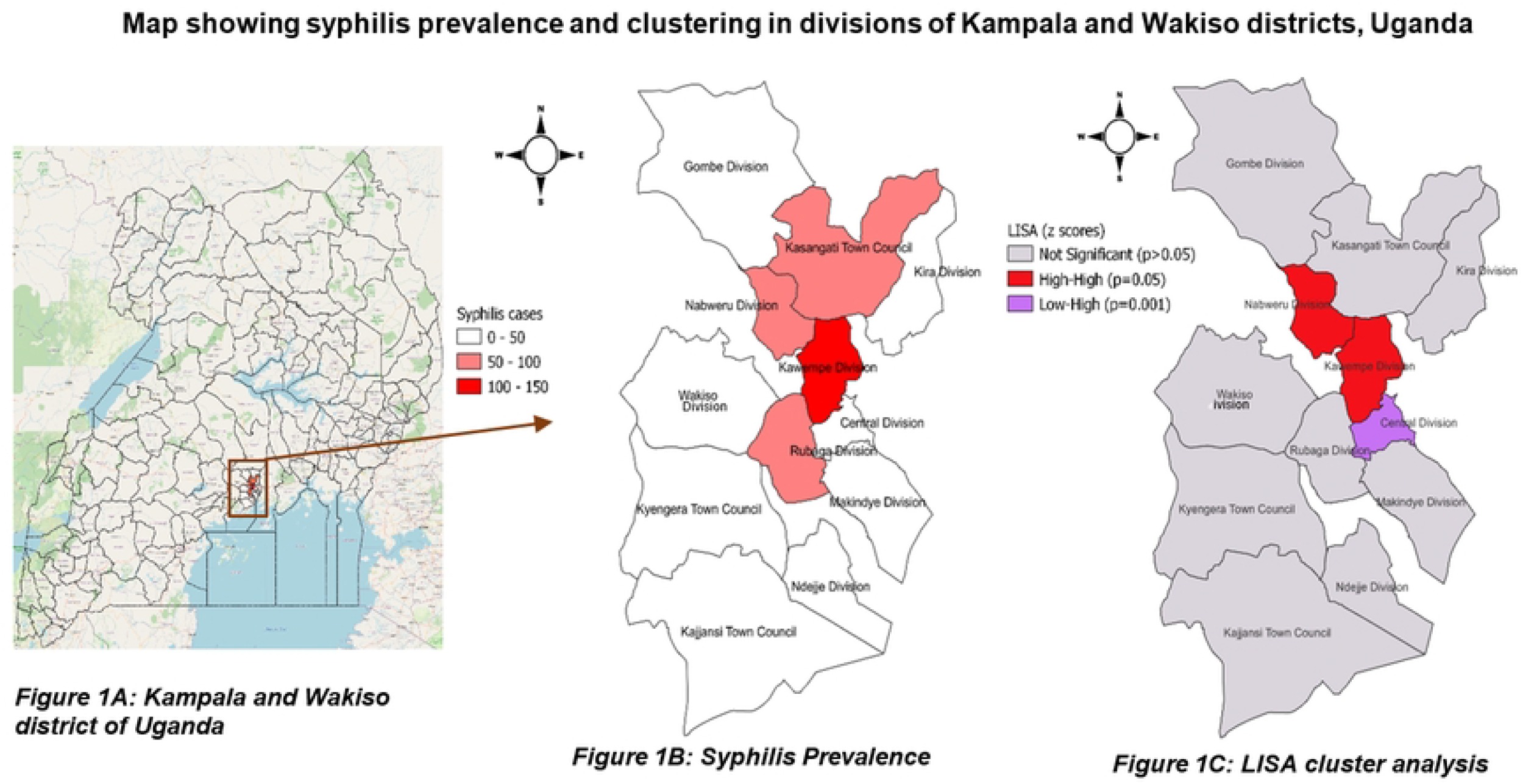
Prevalence and clustering of syphilis in divisions of Kampala and Wakiso listricts, Uganda

Kulldorff modelling showed that divisions with the highest relative risk were Kasangati Town Council (Relative Risk [RR]=2.79, p<0.0001), Kawempe (RR=2.52, p<0.0001), Nabweru (RR=1.95, p=0.0002), and Kyengera Town Council (RR=0.12, p<0.0001) (**Table 4**). Nine (75%) of the 12 divisions had at least one woman living with HIV; however, HIV clustering was not statistically significant across any of the divisions (p>0.05).

**Table 4:** Kulldorff spatial analysis showing distribution of syphilis cases in statistically significant high prevalence divisions.

| Division | AGYW population | Cases | Expected cases | Log-likelihood ratio | RR | P-value |
| --- | --- | --- | --- | --- | --- | --- |
| Kasangati Town Council | 656 | 81 | 33.06 | 27.7 | 2.79 | <0.0001 |
| Kawempe Division | 1267 | 131 | 63.84 | 33.63 | 2.52 | <0.0001 |
| Nabweru Division | 597 | 55 | 30.08 | 9.07 | 1.95 | 0.0002 |
| Kyengera Town Council | 909 | 6 | 45.8 | 29.62 | 0.12 | <0.0001 |

## DISCUSSION

This study of young women living with syphilis in the central districts of Uganda (Kampala and Wakiso) found that syphilis was clustered in divisions with high syphilis prevalence, and divisions neighbouring those with high prevalence. Syphilis prevalence clustering was associated with being in a polygamous relationship, having prior unintended pregnancy, and having tested for HIV more than three months prior.

We identified high syphilis prevalence among young women residing in Kawempe, Nabweru and Rubaga divisions and Kasangati Town Council. Across all divisions, global Moran’s I detected random patterns of syphilis prevalence. However, from the individual analysis of Local Moran’s I, clustering was detected within some hot spots, such as Kawempe and Nabweru divisions. Furthermore, LISA analysis revealed spatial clustering in certain divisions, notably Nabweru and Kawempe, where the prevalence was significantly higher (high-high). Bwaise, one of Kampala’s biggest slums, is located in Kawempe division, and has an HIV prevalence of 25.4% (five times the national prevalence), due to transactional sex, drug/alcohol use, poverty and inconsistent condom use in bar settings [19]. Both Kawempe and Nabweru divisions have large populations, informal economy, and brothels, factors found in previous studies across sub-Saharan Africa to influence sexual behaviour and STI incidence [20, 21].

On the other hand, the Central Division of Kampala District had a lower prevalence despite being surrounded by higher-prevalence divisions. This could be because the Central Division is primarily commercial, has the smallest population compared to other Kampala city divisions and a high concentration of health facilities enabling better access to health services. A previous study in South Africa highlighted a similar pattern in clusters of STI prevalence; areas with better access to preventive services had significantly lower STI prevalence than their neighbouring districts [22]. Understanding these protective factors could inform strategies for controlling syphilis in neighbouring high-prevalence areas. Overall, spatial analysis of syphilis prevalence detailed the importance of geographically targeted interventions, as highlighted in similar STI research in South Africa [22]. In our analyses, one-third were HP divisions, with the number of syphilis cases almost twice that of LP divisions, which was evidence for pockets of syphilis prevalence. Syphilis risk was highest in Kasangati Town Council, followed by Kawempe and Nabweru divisions, and Kyengera Town Council. These divisions share several socio-demographic, geographic, and infrastructural characteristics that would explain the clustering of STIs like syphilis. They are peri-urban or urbanizing areas characterized by high population density, large informal settlements, and limited access to consistent, high-quality healthcare [23]. A systematic review of 23 studies in sub-Saharan Africa revealed that peri-urban areas often serve as hotspots for STIs due to rapid urbanization, weak health systems, and socio-economic vulnerability [24]. HP divisions in our study had dense populations with high levels of informal economic activity and a young demographic, factors associated with increased sexual risk behaviours and limited access to youth-friendly sexual and reproductive health services [23, 25]. They are located along key transport routes, where higher mobility, economic insecurity, and transactional sex are commonly reported—conditions shown to drive STI transmission in urban communities [26, 27].

We observed similar numbers of women in polygamous relations in both HP and LP clusters. In the HP clusters, more women had had an HIV test in the prior three months and were aware of their HIV status. Women who were in polygamous relations, or those who had an unintended pregnancy, or who had an HIV test over three months before the study, were less likely to reside in HP divisions. Such inverse relationships were observed in previous studies [28, 29] which indicated that in more stable, low-transmission communities, polygamous relationships were accompanied by structured social norms and better partner communication, potentially fostering increased health-seeking behaviour. Similarly, women who had experienced unintended pregnancies could have interacted more frequently with maternal and reproductive health services [30, 31]. In addition, the fact that women who had last tested for HIV more than three months prior were more likely to be in LP divisions may reflect a lower perceived or actual risk of recent exposure in these areas.

In Uganda, the social context in which the AGYW are embedded may also play a role in influencing their behaviours and health outcomes. By contrast, socially and economically disadvantaged communities may offer few sexual reproductive resources to help AGYW develop physical, social, and emotional competencies necessary for reaching their full potential for health and well-being. Therefore, if real-level contextual factors contribute to sexual behaviour, thus influencing the syphilis prevalence, then differential prevalence should be expected within large geographical areas such as divisions in which variations in these same factors are present.

This study had several strengths. First, the application of spatial epidemiological methods—including LISA and Kulldorff’s spatial scan statistics—enabled precise identification of syphilis hotspots, providing granular, location-specific insights that are critical for targeted interventions. Such methodological approaches align with global best practices in STI surveillance and support the use of geospatial intelligence in public health decision-making [32, 33]. The study focused on urban and peri-urban areas within the Kampala-Wakiso corridors, thus offering a valuable contribution to understanding intra-urban inequalities in sexual health outcomes, a growing area of interest in rapidly urbanizing low- and middle-income countries. Integrating individual-level behavioural and socio-demographic data with area-level clustering further enriched the analysis, allowing for a nuanced exploration of the interplay between personal risk factors and spatial vulnerability.

Our analysis had limitations. First, the available data was obtained from a single site (Mulago Hospital); although the site had a large catchment area, it presents potential bias in the data. The young women’s population size data per division was also unavailable. Hence, we extracted the population size using previous survey reports. The available data did not include sexual risk behaviour and alcohol use, yet they may influence or confound the associations with syphilis/HIV clustering. Finally, the proportion of young women at risk of syphilis/HIV infection was unknown because, general population surveys did not disaggregate data for pregnant young women. We assumed the proportion of the young women population at greatest risk because we wanted to be modest in our estimation.

## CONCLUSIONS

Our analyses provide information on finer geographic areas of focus in developing syphilis prevention and treatment activities, facilitating equitable resource allocation across regions and subnational units. Priority ought to be given to pregnant women who present with a history of undesired pregnancy, being in a polygamous relationship, and having had an HIV test beyond 3 months. Therefore, STI/HIV programs in this setting may need to rethink the targeting of interventions based on specific individual characteristics or target populations.

## Data Availability

The dataset is publicly available on the repository with this DOI: https://doi.org/10.5281/zenodo.16874894.

## Author contributions

RN and AK designed the study. RN wrote the first draft along with AM and AK. RN performed the data management and statistical analyses. All authors contributed to interpreting the results, writing the manuscript, and approving the final draft.

## Acknowledgements

We are grateful to the Principal Investigators of grant NCT02262390 and the participants for their participation and dedication. We also thank the study team members at the research site for their contributions to data collection. Since participants provided consent to STOP trial participation as demonstrated in the study’s primary paper [14], Human Ethics and Consent to Participate declaration was not applicable for this secondary data analysis.

